# PROGNOSTIC FACTORS FOR CLINICAL COURSE OF PATIENTS WITH COVID-19: PROTOCOL FOR A RAPID LIVING SYSTEMATIC REVIEW

**DOI:** 10.1101/2020.05.06.20087692

**Authors:** César Ramos Rocha-Filho, Ana Carolina Pereira Nunes Pinto, Aline Pereira da Rocha, Keilla Martins Milby, Felipe Sebastião de Assis Reis, Vinicius Tassoni Civile, Nelson Carvas Junior, Rodolfo Rodrigo Pereira Santos, Gabriel Sodré Ramalho, Giulia Fernandes Moça Trevisani, Laura Jantsch Ferla, Maria Eduarda dos Santos Puga, Virgínia Fernandes Moça Trevisani, Alvaro Nagib Atallah

**Author notes:** Research developed at the Brazilian Cochrane Center.

## Abstract

**CONTEXT AND OBJECTIVE:** Determining prognostic factors in a context of health crises such as the COVID-19 scenario may provide the best possible care for patients and optimize the management and the resource utilization of the health system. Thus, we aim to systematically review the prognostic factors for different outcomes of patients with COVID-19. DESIGN AND SETTING: Protocol for a rapid living systematic review methodology following the recommendations proposed by the Cochrane Handbook. METHODS: We will include cohorts and case-control studies. We will search Medline via PubMed, Embase via Elsevier, Cochrane Library - Cochrane Central Register of Controlled Trials (CENTRAL), Portal Regional BVS-LILACS, Scopus and WebofScience to identify studies. No language restrictions will be applied. We will perform the critical appraisal of included studies with the Quality in Prognosis Studies (QUIPS) tool and the certainty of evidence will be evaluated using the Grading of Recommendations Assessment, Development and Evaluation (GRADE).

## INTRODUCTION

Since the first report issued by the World Health Organization (WHO) in early December 2019, the Coronavirus Disease 2019 (COVID-19) outbreak has escalated rapidly (1). Up to April 29, 2020, more than 3 million cases of severe acute respiratory syndrome coronavirus-2 (SARS-CoV-2) infection had been reported from 210 countries and territories; more than 200 thousand people had died (2).

In the scenario of a Public Health Emergency of International Concern, the spectrum of illness presentation or its severity profile is one of the most important parameters for an effective decision-making (3). It helps medical staff during the assessment of patients when the allocation of limited healthcare resources is a reality. It also helps within the effort to provide the best possible care for patients while ensuring the sustainability of the health system (1, 3).

Prognostic factors are known as good indicators to predict disease progression and severity level (3). Currently, it is well established that the case fatality rate for SARS-CoV-2 infection increases with age and number of comorbidities (1). Other factors, such as the decline of the immune function (4), proinflammatory profile (5) and alterations in the Angiotensin I Converting Enzyme 2 (ACE2) (4) are also being described by relevant clinical reports as predictors for COVID-19 progression.

Despite the promising data, to the best of our knowledge the predictive capacity and reliability of these potential indicators has not been properly clarified yet. Thus, the purpose of this rapid living systematic review is to identify the evidence about prognostic factors in patients with COVID-19, considering the following research questions:

1. In people without the disease, what are the risk and protective factors for SARS-CoV-2 infection?
2. In people with SARS-CoV-2 infection, what are the prognostic factors for hospital admission in symptomatic and asymptomatic patients?
3. What are the risk factors for Intensive Care Unit (ICU) admission and/or respiratory support for adult inpatients with COVID-19?
4. In severe patients in ICUs what are the differences between survivors and non-survivors?

## METHODS

The protocol of this systematic review was registred in the Prospective Register of Systematic Reviews (PROSPERO) platform (CRD42020183437). To conduct the rapid living systematic review, we will employ abbreviated systematic review methods. Compared with the methods of a systematic review, the review team will apply the following methodological shortcuts (6):

– no specific searches of grey literature;
– no independent screen of abstracts.

### Eligibility Criteria

#### Types of studies

We will include prospective and retrospective longitudinal cohorts. In view of the limited amount of information, we will also include case-control studies. We will not include cross-sectional studies, as it is not possible to determine prognosis from this design.

#### Types of participants

We will include studies that have evaluated patients with confirmed diagnosis of infection of SARS-CoV-2.

#### Report characteristics

We will include studies performed since November 2019. No language restrictions will be used in the selection.

### Data Sources and Searches

We will search MEDLINE via PubMed, EMBASE via Elsevier, Cochrane Library - Cochrane Central Register of Controlled Trials (CENTRAL), Scopus, Portal Regional BVS - LILACS, and Webof Science, using relevant descriptors and synonyms, adapting the search to the requirements of each database. We will also search in the World Health Organization International Clinical Trials Registry Platform (WHO ICTRP) and ClinicalTrials.gov aiming to identify published, ongoing, and unpublished studies. Finally, we will use the technique of snowballing, searching the lists of references of the included studies.

### Search Strategy

We will use the terms related to the problem of interest and the filter for prognostic studies provided by Wilczynski and Haynes (8). The search strategy in MEDLINE via Pubmed is shown in Table 1.

**Table 1.** Systematic review Search Strategy

| Number | Combiners | Terms |
| --- | --- | --- |
| 1 | Problem of interest | "COVID-19" [Supplementary Concept OR (COVID 19) OR (COVID-19) OR (2019-nCoV) OR (nCoV) OR (Covid19) OR (SARS-CoV) OR (SARSCov2 or ncov*) OR (SARSCov2) OR (2019 coronavirus*) OR (2019 corona virus*) OR (Coronavirus (COVID-19)) OR (2019 novel coronavirus disease) OR (COVID-19 pandemic) OR (COVID-19 virus infection) OR (coronavirus disease-19) OR (2019 novel coronavirus infection) OR (2019-nCoV infection) OR (coronavirus disease 2019) OR (2019-nCoV disease) OR (COVID-19 virus disease) OR "severe acute respiratory syndrome coronavirus 2" [Supplementary Concept] OR (Wuhan coronavirus) OR (Wuhan seafood market pneumonia virus) OR (COVID19 virus) OR (COVID-19 virus) OR (coronavirus disease 2019 virus) OR (SARS-CoV-2) OR (SARS2) OR (2019-nCoV) OR (2019 novel coronavirus) |
| 2 | Type of Study | ((incidence[MeSH:noexp] OR mortality[MeSH Terms] OR follow up studies[MeSH:noexp] OR prognos*[Text Word] OR predict*[Text Word] OR course*[Text Word])<br>#1 AND #2 AND #3 |
| 3 | Filters: | **Publication date from 2019/11/01** |

The search strategy above will be used in Medline via Pubmed and will be adapted to the specifications of each database.

### Study Selection

Based on pre-specified eligibility criteria, two authors will select the studies for inclusion in the review (KMM and ACPNP). When duplicated studies are found in more than one database (studies using the same participants and different outcome measurements or using different time points for the assessments), both reports will be included, but the two reports will be considered as parts of only one study. When duplicated reports are found, e.g. studies with the same participants, with the same outcome measurements and using the same time points for the assessments, the report with the smaller sample size will be excluded. After removing duplicate studies and reports, the authors will read the study titles and abstracts. Studies that clearly do not match the inclusion criteria for the review will be excluded. The selected studies will then be fully read for further scrutiny. The reasons for exclusion of the studies that are fully read will be presented. Disagreements between authors regarding study inclusion will be resolved by the third author (APR). To optimize the process of screening and selection of studies, we will use Rayyan application (9).

### Data Extraction and management

Two authors (ACPNP and CRRF) will independently extract data. Discrepancies or disagreements will be solved by a third author (APR). A predefined form will be used to extract data from included studies. The form will include the following information: (i) the patients (demographic and clinical characteristics); (ii) setting; (iii) time points used for the assessments; (iv) - number of patients lost to follow-up (in each group); (v) reasons for loss to follow up; (vi) - interventions being received; (vii) approach for handling missing data (data imputation/how data imputation was performed, use of intention-to-treat approach); (viii) sources of funding; (ix) possibility of conflict of interests; (x) adverse events; (xi) outcome measures; (xii) protocol deviations.

To assess the feasibility of performing a meta-analysis, we will also extract data for each primary and secondary outcome measure: (i) total number of patients (in each group); (ii) number of events in each group (for dichotomous outcomes);(iii) mean, standard deviation, standard error, median, interquartile range, minimum, maximum, 95% confidence interval (CI) (for continuous outcomes); (iv) p value; (v) hazard ratios with their respective standard errors or confidence intervals (95%).

### Assessment of Methodological Quality in included studies and certainty of evidence

We will perform critical appraisal of included studies with Quality in Prognosis Studies (QUIPS) (10) scale as recommended by Cochrane Collaboration. We will evaluate the certainty of evidence using the Grading of Recommendations Assessment, Development and Evaluation (GRADE) (11). GRADE judgement is based on the overall risk of bias, consistency of the results, directness of the evidence, publication bias and precision of the results for each outcome. The GRADE profiler software, available online, will be used to summarize our findings on the quality of evidence (12). Assessment of risk of bias (ACPNP and APR), and assessment of the quality of evidence (VTC and NCJ) will be performed by two review authors. All the disagreements in the assessment of the risk of bias or quality of evidence will be solved through discussion or, if required, by consulting with a third author (ANA).

### Data Synthesis and Analysis

We will perform analyses according to the recommendations of Cochrane, and the Cochrane Prognosis Methods Group, and we will use Review Manager 5 to perform meta-analysis when possible for hazard or odds ratios.

We will pool hazard ratios (unadjusted (crude) or adjusted) or odds ratio with their standard errors for hospital admission, intensive care unit admission and/or respiratory support for adult inpatients with COVID-19 and mortality, using generic inverse variance method with random-effects model.

We will also pool incidence results (for prognostic factors) with their respective confidence intervals (95%) by the inverse variance method with a random-effects model, using the DerSimonian-Laird estimator for τ2. We will adjust data by Freeman-Tukey double arcosen transformation and confidence intervals will be calculate by the Clopper-Pearson method for individual studies. For these data, we will perform proportion meta-analysis using RStudio^©^ software, with the “meta” package (version 4.9-6) and “metaprop” function.

### Dealing with missing data

For studies that do not provide an HR and associated standard error (SE), we will use information and results reported in the text, tables, and Kaplan-Meier (K-M) curves. We will contact the principal investigators of included studies asking for additional data, or to clarify issues regarding the studies. In the absence of a reply from the authors, we will expose the data in a descriptive manner avoiding imputation.

### Assessment of heterogeneity

We will employ the Cochran’s Q test to assess the presence of heterogeneity considering a threshold of P value < 0.1 as an indicator of whether heterogeneity is present.

In addition, we will assess statistical heterogeneity by examining the Higgins I^2^ statistic following these thresholds:

· < 25%: no (none) heterogeneity;
· 25% to 49%: low heterogeneity;
· 50% to 74%: moderate heterogeneity;
· ≥ 75%: high heterogeneity.

## DISCUSSION

This rapid living review will systematically evaluate the best available evidence to identify the risk and protective factors of COVID-19, which we expect will help the front line on their decision making processes. While some data (13) have shown older adults are at higher risk for worse outcomes, other studies have raised additional questions on whether factors such as the decline in immune function (4), proinflammatory profile (5) and alterations on the angiotensin I converting enzyme 2 (ACE2) (4) could also constitute risk factors for worse outcomes. At the same time, our review aims to clarify the uncertainty over which characteristics could constitute protective factors once a person has been exposed to SARSCoV-2.

To ensure the quality of the results, we will follow the Cochrane Handbook of Systematic Reviews recommendations (7). If possible, we plan to pool data into meta-analysis for reducing the probability of type 2 error within the comparisons. Potential limitations for this study include the possibility of finding biased studies which can make them unsuitable for clustering or meta-analysis, or small sample studies that do not allow us to provide precise estimates.

We believe that the strengths of this rapid systematic review include the transparency, the strict methods, the evaluation of the quality of evidence, and the extensive and more sensitive searches. This being said, we will be able to identify the current available evidence, differentiating the prognostic factors for each stage of the disease and to provide important information for clinical decision-making on coronavirus disease 2019 (COVID-19) that has recently emerged and caused a deadly pandemic.

### Reporting standards

This systematic review protocol was written as per the PRISMA-P guidelines (14).

## Data Availability

The authors confirm that the data supporting the findings of this study are available within the article

## APPENDIX

#### Cochrane Library

#1 MeSH descriptor: [Coronavirus] explode all trees
#2 MeSH descriptor: [Coronaviridae] explode all trees
#3 MeSH descriptor: [Betacoronavirus] explode all trees
#4 MeSH descriptor: [Coronavirus Infections] explode all trees
#5 (COVID 19) OR (COVID-19) OR (2019-nCoV) OR (nCoV) OR (Covid19) OR (SARS-CoV) OR (SARSCov2 or ncov*) OR (SARSCov2) OR (2019 coronavirus*) OR (2019 corona virus*) OR (Coronavirus (COVID-19)) OR (2019 novel coronavirus disease) OR (COVID-19 pandemic) OR (COVID-19 virus infection) OR (coronavirus disease-19) OR (2019 novel coronavirus infection) OR (2019-nCoV infection) OR (coronavirus disease 2019) OR (2019-nCoV disease) OR (COVID-19 virus disease) OR “severe acute respiratory syndrome coronavirus 2” [Supplementary Concept] OR (Wuhan coronavirus) OR (Wuhan seafood market pneumonia virus) OR (COVID19 virus) OR (COVID-19 virus) OR (coronavirus disease 2019 virus) OR (SARS-CoV-2) OR (SARS2) OR (2019-nCoV) OR (2019 novel coronavirus)
#6 #1 OR #2 OR #3 OR #4 OR #5

#### EMBASE

#1 ‘covid 19’/exp OR (COVID 19) OR (COVID-19) OR (2019-nCoV) OR (nCoV) OR (Covid19) OR (SARS-CoV) OR (SARSCov2 or ncov*) OR (SARSCov2) OR (2019 coronavirus*) OR (2019 corona virus*) OR (Coronavirus (COVID-19)) OR (2019 novel coronavirus disease) OR (COVID-19 pandemic) OR (COVID-19 virus infection) OR (coronavirus disease-19) OR (2019 novel coronavirus infection) OR (2019-nCoV infection) OR (coronavirus disease 2019) OR (2019-nCoV disease) OR (COVID-19 virus disease) OR (severe acute respiratory syndrome coronavirus 2) OR (Wuhan coronavirus) OR (Wuhan seafood market pneumonia virus) OR (COVID19 virus) OR (COVID-19 virus) OR (coronavirus disease 2019 virus) OR (SARS-CoV-2) OR (SARS2) OR (2019-nCoV) OR (2019 novel coronavirus)
#2 #1 AND [embase]/lim NOT ([embase]/lim AND [medline]/lim)

#### PORTAL REGIONAL BVS - LILACS

MH:”Infecções por Coronavirus” OR (Infecções por Coronavirus) OR (Infecciones por Coronavirus) OR (Coronavirus Infections) OR (COVID-19) OR (COVID 19) OR (Doença pelo Novo Coronavírus (2019-nCoV)) OR (Doença por Coronavírus 2019nCoV) OR (Doença por Novo Coronavírus (2019-nCoV)) OR (Epidemia de Pneumonia por Coronavirus de Wuhan) OR (Epidemia de Pneumonia por Coronavírus de Wuhan) OR (Epidemia de Pneumonia por Coronavírus de Wuhan de 2019-2020) OR (Epidemia de Pneumonia por Coronavírus em Wuhan) OR (Epidemia de Pneumonia por Coronavírus em Wuhan de 2019-2020) OR (Epidemia de Pneumonia por Novo Coronavírus de 2019-2020) OR (Epidemia pelo Coronavírus de Wuhan) OR (Epidemia pelo Coronavírus em Wuhan) OR (Epidemia pelo Novo Coronavírus (2019-nCoV)) OR (Epidemia pelo Novo Coronavírus 2019) OR (Epidemia por 2019-nCoV) OR (Epidemia por Coronavírus de Wuhan) OR (Epidemia por Coronavírus em Wuhan) OR (Epidemia por Novo Coronavírus (2019-nCoV)) OR (Epidemia por Novo Coronavírus 2019) OR (Febre de Pneumonia por Coronavírus de Wuhan) OR (Infecção pelo Coronavírus 2019-nCoV) OR (Infecção pelo Coronavírus de Wuhan) OR (Infecção por Coronavirus 2019-nCoV) OR (Infecção por Coronavírus 2019-nCoV) OR (Infecção por Coronavírus de Wuhan) OR (Infecções por Coronavírus) OR (Pneumonia do Mercado de Frutos do Mar de Wuhan) OR (Pneumonia no Mercado de Frutos do Mar de Wuhan) OR (Pneumonia por Coronavírus de Wuhan) OR (Pneumonia por Novo Coronavírus de 2019-2020) OR (Surto de Coronavírus de Wuhan) OR (Surto de Pneumonia da China 2019-2020) OR (Surto de Pneumonia na China 2019-2020) OR (Surto pelo Coronavírus 2019-nCoV) OR (Surto pelo Coronavírus de Wuhan) OR (Surto pelo Coronavírus de Wuhan de 2019-2020) OR (Surto pelo Novo Coronavírus (2019-nCoV)) OR (Surto pelo Novo Coronavírus 2019) OR (Surto por 2019-nCoV) OR (Surto por Coronavírus 2019nCoV) OR (Surto por Coronavírus de Wuhan) OR (Surto por Coronavírus de Wuhan de 2019-2020) OR (Surto por Novo Coronavírus (2019-nCoV)) OR (Surto por Novo Coronavírus 2019) OR (Síndrome Respiratória do Oriente Médio) OR (Síndrome Respiratória do Oriente Médio (MERS)) OR (Síndrome Respiratória do Oriente Médio (MERS-CoV)) OR (Síndrome Respiratória do Oriente Médio por Coronavírus) OR MH:<u>C01.925.782.600.550.200</u>$

#### SCOPUS

#1 TITLE-ABS-KEY(coronavirus)
#2 TITLE-ABS-KEY(coronaviridae)
#3 TITLE-ABS-KEY(“Coronavirus Infections”)
#4 TITLE-ABS-KEY(betacoronavirus)
#5 (COVID 19) OR (COVID-19) OR (2019-nCoV) OR (nCoV) OR (Covid19) OR (SARS-CoV) OR (SARSCov2 or ncov*) OR (SARSCov2) OR (2019 coronavirus*) OR (2019 corona virus*) OR (Coronavirus (COVID-19)) OR (2019 novel coronavirus disease) OR (COVID-19 pandemic) OR (COVID-19 virus infection) OR (coronavirus disease-19) OR (2019 novel coronavirus infection) OR (2019-nCoV infection) OR (coronavirus disease 2019) OR (2019-nCoV disease) OR (COVID-19 virus disease) OR (severe acute respiratory syndrome coronavirus 2) OR (Wuhan coronavirus) OR (Wuhan seafood market pneumonia virus) OR (COVID19 virus) OR (COVID-19 virus) OR (coronavirus disease 2019 virus) OR (SARS-CoV-2) OR (SARS2) OR (2019nCoV) OR (2019 novel coronavirus)
#6 #1 OR #2 OR #3 OR #4 OR #5
#7 #7 AND #6

#### WEB OF SCIENCE

#1 (COVID 19) OR (COVID-19) OR (2019-nCoV) OR (nCoV) OR (Covid19) OR (SARS-CoV) OR (SARSCov2 or ncov*) OR (SARSCov2) OR (2019 coronavirus*) OR (2019 corona virus*) OR (Coronavirus (COVID-19)) OR (2019 novel coronavirus disease) OR (COVID-19 pandemic) OR (COVID-19 virus infection) OR (coronavirus disease-19) OR (2019 novel coronavirus infection) OR (2019-nCoV infection) OR (coronavirus disease 2019) OR (2019-nCoV disease) OR (COVID-19 virus disease) OR (severe acute respiratory syndrome coronavirus 2) OR (Wuhan coronavirus) OR (Wuhan seafood market pneumonia virus) OR (COVID19 virus) OR (COVID-19 virus) OR (coronavirus disease 2019 virus) OR (SARS-CoV-2) OR (SARS2) OR (2019nCoV) OR (2019 novel coronavirus)

## REFERENCES

1. Beeching NJ, Fletcher TE, Fowler R. Coronavirus disease 2019 (COVID-19) BMJ Best Practice. 2020.

2. World Health Organization. Coronavirus disease 2019 (COVID-19). Situation Report - 100. World Health Organization [Internet]. 2020. Available from: https://www.who.int/docs/default-source/coronaviruse/situation-reports/20200429-sitrep-100covid-19.pdf?sfvrsn=bbfbf3d1_2

3. Wu JT, Leung K, Bushman M, Kishore N, Niehus R, de Salazar PM, et al. Estimating clinical severity of COVID-19 from the transmission dynamics in Wuhan, China. Nature Medicine. 2020;26(4):506–10. DOI: 10.1038/s41591-020-0822-7.

4. Morley, J.E., Vellas, B. COVID-19 and Older Adult. J Nutr Health Aging 24, 364–365 (2020). https://doi.org/10.1007/s12603-020-1349-9

5. Lakatta EG. The reality of getting old. Nat Rev Cardiol. 2018;15(9):499–500. DOI: 10.1038/s41569-018-0068-y.

6. Garritty C, Gartlehner G, Kamel C, King VJ, Nussbaumer-Streit B, Stevens A, et al. Cochrane Rapid Reviews Interim Guidance from the Cochrane Rapid Reviews Methods Group 2020. Available from: https://methods.cochrane.org/rapidreviews/sites/methods.cochrane.org.rapidreviews/files/public/uploads/cochrane_rr_-_guidance-23mar2020-v1.pdf.

7. Higgins J. Cochrane Handbook for Systematic Reviews of Interventions 2011.

8. Wilczynski NL, Haynes RB for the Hedges Team. Developing optimal search strategies for detecting clinically sound prognostic studies in MEDLINE: an analytic survey. BMC Med. 2004 Jun 09;2(1):23

9. Ouzzani M, Hammady H, Fedorowicz Z, Elmagarmid A. Rayyan-a web and mobile app for systematic reviews. Syst Rev. 2016;5(1):210. DOI: 10.1186/s13643-016-0384-4.

10. Hayden J, van der Windt D, Cartwright J, Côté P, Bombardier C. Assessing Bias in Studies of Prognostic Factors. Annals of internal medicine. 2013;158:280–6. DOI: 10.7326/0003-4819-158-4-201302190-00009.

11. Atkins D, Best D, Briss PA, Eccles M, Falck-Ytter Y, Flottorp S, et al. Grading quality of evidence and strength of recommendations. Bmj. 2004;328(7454):1490. DOI: 10.1136/bmj.328.7454.1490.

12. GDT G. Grade’s software for summary of findings tables, health technology assessment and guidelines 2015 [Available from: https://gradepro.org/.

13. Wu Z, McGoogan JM. Characteristics of and Important Lessons From the Coronavirus Disease 2019 (COVID-19) Outbreak in China: Summary of a Report of 72–314 Cases From the Chinese Center for Disease Control and Prevention. JAMA. 2020;323(13):1239–42. DOI: 10.1001/jama.2020.2648.

14. Moher D, Shamseer L, Clarke M, Ghersi D, Liberati A, Petticrew M, et al. Preferred reporting items for systematic review and meta-analysis protocols (PRISMA-P) 2015 statement. Syst Rev. 2015;4:1. DOI: 10.1186/2046-4053-4-1.

